# Use of Electronic Health Records to Characterize Patients with Uncontrolled Hypertension in Two Large Health System Networks

**DOI:** 10.1101/2023.07.26.23293225

**Authors:** Yuan Lu, Ellen C. Keeley, Eric Barrette, Rhonda M. Cooper-DeHoff, Sanket S. Dhruva, Jenny Gaffney, Ginger Gamble, Bonnie Handke, Chenxi Huang, Harlan M. Krumholz, Caitrin W McDonough Rowe, Wade Schulz, Kathryn Shaw, Myra Smith, Jennifer Woodard, Patrick Young, Keondae Ervin, Joseph S. Ross

**Author notes:** **Address correspondence to:** Joseph S. Ross, MD, MHS, 367 Cedar Street, Ste 405B, New Haven, CT 06510, Yuan Lu, ScD, 195 Church Street, 5^th^ floor.

## Abstract

**Background:** Improving hypertension control is a public health priority. However, uncertainty remains regarding the optimal way to identify patients with uncontrolled hypertension using electronic health records (EHR) data.

**Methods:** In this retrospective cohort study, we applied computable definitions to the EHR data to identify patients with controlled and uncontrolled hypertension and to evaluate differences in characteristics, treatment, and clinical outcomes between these patient populations. We included adult patients (≥18 years) with hypertension receiving ambulatory care within Yale-New Haven Health System (YNHHS; a large US health system) and OneFlorida Clinical Research Consortium (OneFlorida; a Clinical Research Network comprised of 16 health systems) between October 2015 and December 2018. We identified patients with controlled and uncontrolled hypertension based on either a single blood pressure (BP) measurement from a randomly selected visit or all BP measurements recorded between hypertension identification and the randomly selected visit).

**Results:** Overall, 253,207 and 182,827 adults at YNHHS and OneFlorida were identified as having hypertension. Of these patients, 83.1% at YNHHS and 76.8% at OneFlorida were identified using ICD-10-CM codes, whereas 16.9% and 23.2%, respectively, were identified using elevated BP measurements (≥ 140/90 mmHg). Uncontrolled hypertension was observed among 32.5% and 43.7% of patients at YNHHS and OneFlorida, respectively. Uncontrolled hypertension was disproportionately higher among Black patients when compared with White patients (38.9% versus 31.5% in YNHHS; p<0.001; 49.7% versus 41.2% in OneFlorida; p<0.001). Medication prescription for hypertension management was more common in patients with uncontrolled hypertension when compared with those with controlled hypertension (overall treatment rate: 39.3% versus 37.3% in YNHHS; p=0.04; 42.2% versus 34.8% in OneFlorida; p<0.001). Patients with controlled and uncontrolled hypertension had similar rates of short-term (at 3 and 6 months) and long-term (at 12 and 24 months) clinical outcomes. The two computable definitions generated consistent results.

**Conclusions:** Computable definitions can be successfully applied to health system EHR data to conduct population surveillance for hypertension and identify patients with uncontrolled hypertension who may benefit from additional treatment.

**Clinical Perspective:** *What is new?:*

- In this retrospective study that included 253,207 and 182,827 hypertensive adults at Yale-New Haven Health System and OneFlorida Clinical Research Consortium, we applied two computable definitions to identify patients with uncontrolled hypertension.
- The two computable definitions generated consistent results and showed that approximately 30-40% of hypertensive patients have uncontrolled hypertension, of whom 60% were untreated or undertreated.

*What are the clinical implications?:*

- Computable definitions can be successfully applied to health system EHR data to conduct population surveillance for hypertension and identify patients with uncontrolled hypertension who may benefit from additional treatment.

## INTRODUCTION

Improving hypertension control is a public health priority in the US.^1^ Approximately half of US adults have hypertension, but fewer than half have their blood pressure (BP) controlled.^2^ Individuals with uncontrolled BP are at high risk for adverse clinical outcomes, including stroke, myocardial infarction, kidney disease, heart failure and cognitive decline.^3^ Therefore, characterizing these individuals, as well as their treatment and outcome patterns, is critical to informing both public health and health system interventions. Electronic health record (EHR) data present new opportunities to better understand uncontrolled hypertension because EHRs provide more efficient access to a wider range of detailed longitudinal clinical information compared with other data sources (e.g., claims databases and clinical registries).^4, 5^

However, uncertainty remains regarding the optimal way to define and identify patients with uncontrolled hypertension using EHR data. There is no specific diagnostic code for uncontrolled hypertension, and a diagnosis usually requires many observations over time. Complex phenotypes like this can benefit from computable definitions that utilize various data elements from the EHR to identify patients with a specific disease.^6-8^ While current clinical guidelines have established a basic definition of uncontrolled hypertension,^3, 9^ few studies have developed computable definitions for uncontrolled hypertension based on structured diagnosis codes, vital signs, and using common data models for use in clinical research and practice. Moreover, EHR data elements can be assembled in multiple ways in terms of frequency, clinical context, and time. Yet, it is unclear how different computable definitions of uncontrolled hypertension may influence patient cohort identification. This information is essential for identifying people with hypertension who would benefit from more aggressive management and will lay the foundation for assessments the quality of care and outcomes of these patients.

Accordingly, we developed and applied two computable definitions to retrospectively identify patients with controlled and uncontrolled hypertension using EHR data from two large health system networks. We also compared characteristics, treatment patterns, and clinical outcomes of patients with controlled and uncontrolled hypertension.

## METHODS

### Project Origination

The National Evaluation System for health Technology Coordinating Center (NESTcc) is an organization established through grant funding to the Medical Device Innovation Consortium by the US Food and Drug Administration in 2016 to promote the development of robust real-world evidence for regulatory decision-making.^10^ NESTcc currently includes 19 Network Collaborators (health care providers, academic research institutions, payers, and professional registries) that collect, curate, and analyze real-world evidence that may be used for regulatory decision-making.

This study was proposed to NESTcc by Medtronic Inc, which is currently studying its Symplicity™ Renal Denervation System in patients with hypertension in a series of sham-controlled and real-world studies intended to support a premarket approval application in the USA.^11, 12^ After an independent review of the study concept and subsequent proposal, NESTcc funded the project. Among its Network Collaborators, NESTcc identified a large health system and a clinical research network interested in pursuing the proposed project, each of which had extensive experience with EHR data analysis: Yale-New Haven Health System (YNHHS) and the OneFlorida Clinical Research Consortium (OneFlorida). Medtronic and the two NESTcc Network Collaborators, with YNHHS serving as the lead, developed a full research plan that was approved by NESTcc. Institutional Review Board approval was obtained at Yale University and University of Florida. The study followed the guidelines for cohort studies, described in the Strengthening the Reporting of Observational Studies in Epidemiology (STROBE) Statement: guidelines for reporting observational studies.

### Data Sources

The data sources for this study consisted of EHR data from YNHHS and OneFlorida. YNHHS is a large academic health system consisting of five distinct hospital delivery networks and associated ambulatory clinics located in Connecticut and Rhode Island. The system provides services for approximately two million patients annually. OneFlorida is a statewide clinical research network including 16 partner health systems providing services for 40% of Florida’s population.

Both YNHHS and OneFlorida conformed data to the National Patient-Centered Clinical Research Network (PCORnet) common data model via extract/transform/load software,^13, 14^ ensuring data elements were standardized and consistent across the two sites. Both sites conducted data quality assessments in a standardized fashion. Data quality was assessed by performing domain value validation checks periodically, assessing for data relevance, reliability, and robustness. Cross-validation was performed on the various data sources to assess for any data gaps and to ensure data completeness. In addition to internal quality checks at each site, the Yale team and the OneFlorida team met regularly to resolve issues regarding the validity and robustness of the results. For this analysis, we used a versioned extract of the PCORnet common data model from October 1, 2015, when International Classification of Diseases-10th Edition-Clinical Modification (ICD-10-CM) diagnosis was introduced, through December 31, 2018.

### Study Population

The study population included adult patients (≥18 years) who met the clinical criteria of hypertension between October 1, 2015 and December 31, 2018 if (1) they had an ICD-10-CM diagnosis code for hypertension (I10, I11, I12, I13, I15, I16) associated with at least one ambulatory visit, or (2) in the absence of a diagnosis, they had at least two elevated BP measurements (systolic BP [SBP] ≥ 140 mmHg or diastolic BP [DBP] ≥ 90 mmHg) recorded in the EHR at two separate ambulatory visits occurring at least one day apart within a 6-month period at any time between October 1, 2015 and December 31, 2018. Numerous studies in the literature have supported the validity of using these methods for identifying patients with hypertension.^15, 16^ We used BP ≥140/90 mmHg as the cutoff for hypertension because this was the definition of hypertension at the time from which most of the data were extracted.^9^

We excluded patients with fewer than 3 months follow-up time, female patients with diagnostic or procedural evidence of pregnancy (ICD-10-CM [Z33, Z34, O80, O82, O00, O01, O02, O03, O04, O07, O08]) and patients receiving dialysis (ICD-10-CM [Z99.2]). We also included only those BP measurements recorded at ambulatory visits, excluding BP measurements from inpatient and emergency department (ED) encounters because BP measurements in those encounters could be elevated due acute conditions. For any visit with multiple BP measurements recorded, the lowest SBP measurement and lowest DBP measurement were used to ascertain hypertension status. We extended our observation period until the end of 2019 to ensure at least 12-month follow-up for patients.

### Definitions of Controlled and Uncontrolled Hypertension

As there are multiple ways in which the EHR data elements are assembled in terms of frequency, clinical context, and time, we tested two different approaches to operationalize the definitions of controlled and uncontrolled hypertension. Specifically, we randomly selected one ambulatory encounter with a BP measurement occurring at least 3 months after hypertension identification and between October 1, 2015 and December 31, 2018 as the index encounter, then applied two approaches to define controlled/uncontrolled hypertension. Our rationale for selecting a random date to minimize selection bias. If we had chosen the most recent encounter as the index date, our sample would have been biased toward patients with shorter follow-up times, making it less likely for them to achieve blood pressure control. Conversely, if we had selected the earliest encounter, our sample would have been biased towards patients with longer follow-up times, offering more opportunity for the patients to achieve blood pressure control (and experience poor clinical outcomes). By randomly selecting a date, we ensured that the follow-up times for our sample would be more balanced overall. In addition, we required patients to have at least 3 months after hypertension identification before being included in the study. This allowed for a sufficient period for treatment to take effect, and it ensured that patients had a fair chance to achieve blood pressure control regardless of when the index date was selected. To ensure accuracy and reliability of the data, we only included encounters where a BP measurement was documented at the time of the visit.

In approach 1, hypertensive patients were considered to have controlled hypertension if more than 50% of their SBP measurements were < 140 mmHg and DBP measurements were < 90 mmHg among the measured BPs on all ambulatory encounters from the identification date up to and including the index encounter. Hypertensive patients were considered to have uncontrolled hypertension if 50% or more of SBPs were ≥ 140 mmHg or DBPs were ≥ 90 mmHg among the measured BPs on all encounters from the identification date up to and including the index encounter (**Figure 1)**. In approach 2, hypertensive patients were considered to have controlled hypertension when both SBP < 140 mmHg and DBP < 90 mmHg at the index encounter. Hypertensive patients were considered to have uncontrolled hypertension when either the SBP was ≥ 140 mmHg or the DBP was ≥ 90 mmHg at the index encounter. Since approach 1 used multiple BP measurements over time, it comprises the primary analysis while approach 2 is the sensitivity analysis. The National Quality Forum BP measure defined control of hypertension based on a BP reading of <140/90 mmHg at the most recent healthcare encounter. This measure is based on a BP reading from a single encounter, which was consistent with approach 2 of the study. We performed two sensitivity analyses to assess the robustness of our results. In the first analysis, we defined controlled hypertension as having more than 50% of SBP measurements below 130 mmHg and DBP measurements below 80 mmHg among all measured BPs recorded during ambulatory encounters, starting from the identification date and continuing up to and including the index encounter. This threshold was chosen based on established clinical guidelines. In the second sensitivity analysis, we employed a different threshold. Here, controlled hypertension was defined as having more than 75% of SBP measurements below 140 mmHg and DBP measurements below 90 mmHg among all measured blood pressures recorded during ambulatory encounters, starting from the identification date and continuing up to and including the index encounter. This threshold aligns with alternative clinical recommendations.

**Figure 1.**
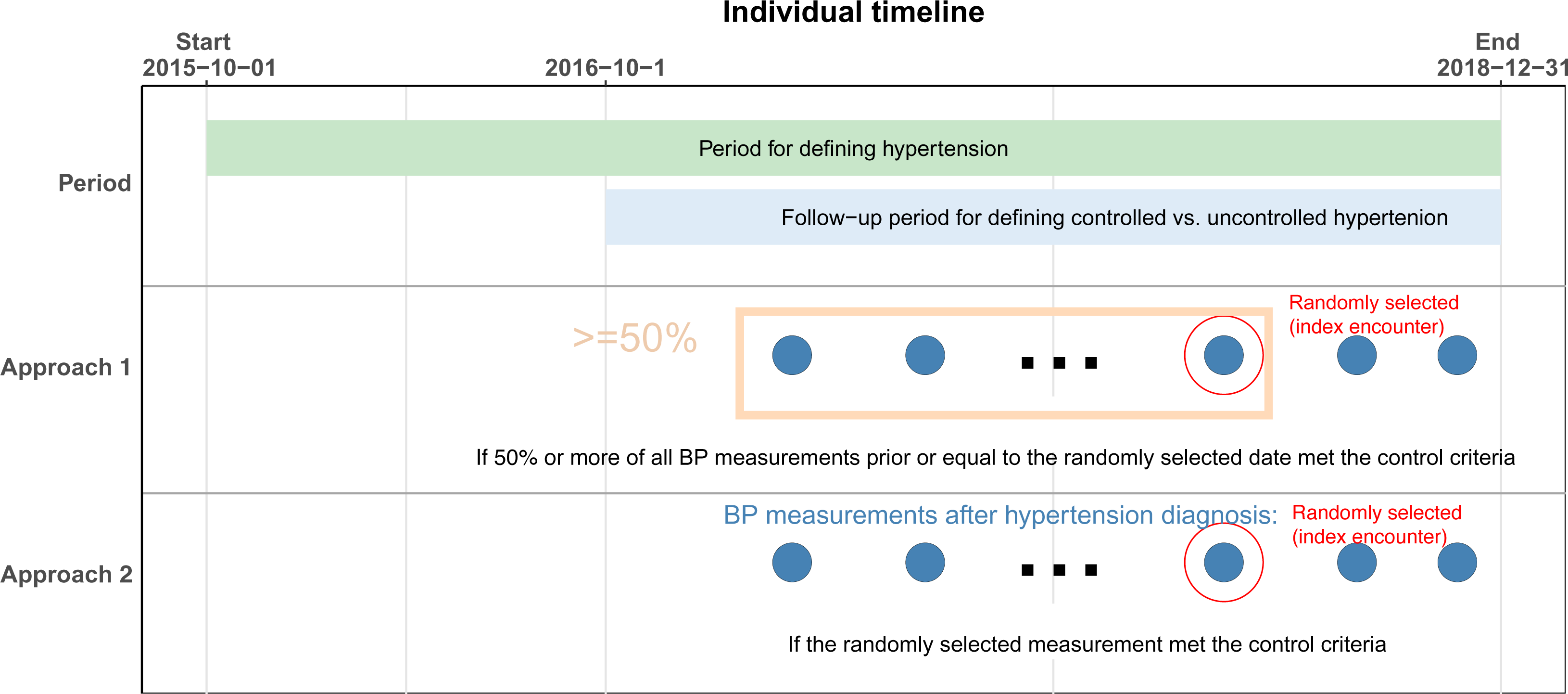
Cohort Definitions for Controlled and Uncontrolled Hypertension. Footnote: The red dot on the graph indicates an ambulatory encounter selected randomly at least three months after hypertension identification and between October 1, 2015, and December 31, 2018, serving as the index encounter. We employed two different approaches to determine controlled hypertension among the hypertensive patients. In approach 1, controlled hypertension was defined as having more than 50% of systolic blood pressure (SBP) measurements below 140 mmHg and diastolic blood pressure (DBP) measurements below 90 mmHg across all ambulatory encounters, from the identification date up to and including the index encounter. In approach 2, controlled hypertension was defined as having both SBP < 140 mmHg and DBP < 90 mmHg at the index encounter.

### Baseline Characteristics

Baseline demographic and clinical characteristics of patients included age, race, ethnicity, sex, health insurance type, smoking status, body mass index [BMI] and comorbidities. Race was categorized as Black, White, other(s), and unknown. Ethnicity was categorized as Hispanic, non-Hispanic, and unknown. Comorbidities included heart failure, diabetes mellitus, history of acute myocardial infarction, coronary artery disease, cerebrovascular disease, stroke, atrial fibrillation or flutter, chronic kidney disease, chronic obstructive pulmonary disease, dyslipidemia, peripheral arterial disease, angina, depression, dementia, hypertensive retinopathy, and substance use disorder.

Characteristics using a set time point such as age were defined based on the index encounter. If data for a specific characteristic were not available from the index encounter (e.g., smoking status), we used the most recent data available prior to the index date. Characteristics such as insurance status, which may change across encounters, were defined based on the index encounter. Comorbidities were defined using ICD-10-CM codes based on the 1-year period prior to the index date (see details in **Supplemental Table S1**).

### Classification of Antihypertensive Medications

To properly classify EHR-based prescription drug data into antihypertensive therapeutic indication and antihypertensive drug classes, we used a previously developed antihypertensive drug classification system based off RxNorm Concept Unique Identifiers (RxCUIs).^17^ We included only oral formulations, with the exception of transdermal clonidine patches. We classified antihypertensive medications into major drug classes, including angiotensin-converting enzyme inhibitors (ACEI), angiotensin receptor blockers (ARB), beta blockers, calcium channel blockers (CCB), thiazide or thiazide-like diuretics, and other antihypertensive drugs. For combination drugs, we classified them into the multiple component classes of the combination drugs. The list of drug ingredient in each antihypertensive drug class was presented in **Supplemental Table S2**.

### Short-term and Long-term Outcomes

We examined pre-specified short-term outcomes at 3 and 6 months and long-term outcomes at 12 and 24 months after the index date. The short-term and long-term outcomes were the same, including clinical outcomes (the composite of death and non-fatal cardiovascular disease [CVD] events) and healthcare utilization (ED visits and hospitalizations for any cause; ambulatory visits for any cause). Non-fatal CVD events were defined as any diagnosis of a specified hypertension-related disease, including acute myocardial infarction (AMI), heart failure, atrial fibrillation/flutter, aortic dissection, renal disease, hemorrhagic stroke, ischemic stroke, or hypertensive crisis at an ED or inpatient visit. Of note, we included only acute event codes, including both primary and secondary diagnosis codes, for outcome ascertainment. We excluded CVD events reported at ambulatory encounters because of the inability to reliably distinguish patients with acute CVD events from those with history of prior CVD. Death was identified through a combination of reported death records in the EHR, a death diagnosis at any visit, and encounters with a discharge status of expired. Social Security Death Master File were also used to identify mortality data. ICD-10-CM diagnosis codes for clinical outcomes are listed in **Supplemental Table S3**.^18^ As longer follow-up periods are likely required to comprehensively assess the complete range of outcomes associated with hypertension, it is important to note that our examination of long-term outcomes at 24 months is conducted as an exploratory analysis within this study.

### Statistical Analyses

We first calculated the prevalence of controlled and uncontrolled hypertension among all patients with hypertension, respectively. We described the demographic and clinical characteristics of the hypertensive population overall and by controlled vs. uncontrolled status. We then described the number and class of antihypertensive medications prescribed both in the year prior to the index date and on the index date among overall hypertensive patients and by controlled vs. uncontrolled status. We also described the three most prescribed antihypertensive medications among patients using 1, 2, and 3 or more antihypertensive medications. Finally, we described the frequency and percentage of patient outcomes and healthcare utilization at 3, 6, 12 and 24 months among overall hypertensive patients and by controlled vs. uncontrolled status. For the analysis of patient characteristics, antihypertensive medication prescriptions, and outcomes at 3, 6, and 12 months, we included individuals with a follow-up period of more than three months but less than 24 months. However, we did not include them in the analysis of outcomes at 24 months due to insufficient follow-up data. To mitigate the concern of potential censoring, we excluded patients from our analysis who had less than 3 months of follow-up time. Moreover, we employed a time-to-event analysis methodology that effectively addressed the variable durations of follow-up among patients when assessing clinical outcomes. Patients were not censored solely due to the absence of documented interactions with the healthcare system at specific time intervals. Instead, their follow-up time was truncated at the most recent recorded visit or appointment in the EHR, ensuring that their data were included up until the last known contact.

Comparisons between uncontrolled and controlled hypertensive patients for characteristics, treatment, and outcomes were performed using appropriate tests, including Pearson’s chi-square test for normally distributed continuous variables, the Wilcoxon signed rank test for non-normally distributed continuous variables, the McNemar test for 2*2 categorical variables and the generalized Mantel-Haenszel test for 2*n categoric variables (where n> 2). All analyses were conducted individually at each site using a decentralized model;^19^ summary results were shared across researchers from the two sites, with no patient-level data shared. All statistical analyses were performed using SAS software version 9.4 (SAS institute, Cary, NC, USA) and Statistical package R version 3.6.

## RESULTS

Overall, 514,687 adult patients from YNHHS and 1,075,204 adult patients from OneFlorida had at least one outpatient visit with BP data recorded between October 1, 2015, and December 31, 2018 (**Figure 2**). Among these patients, 253,207 from YNHHS and 182,827 from OneFlorida had hypertension based on either diagnosis codes or BP elevations and met specific inclusion and exclusion criteria. Of patients with hypertension, 83.1% at YNHHS and 76.8% at OneFlorida were identified based on ICD-10-CM codes, and 16.9% at YNHHS and 23.2% at OneFlorida were identified using elevated BP measures (**Supplemental Table S4**). At YNHHS, the mean age of patients was 65.0 years (SD = 14.6) years and 47.8% of patients were men; 12.6% of patients were Black, 76.2% were White, and 9.0% were Hispanic. At OneFlorida, the mean age of patients was 61.0 years (SD: 14.7) years and 44.8% of patients were men; 25.2% of patients were Black, 47.7% were White, and 15.4% were Hispanic.

**Figure 2.**
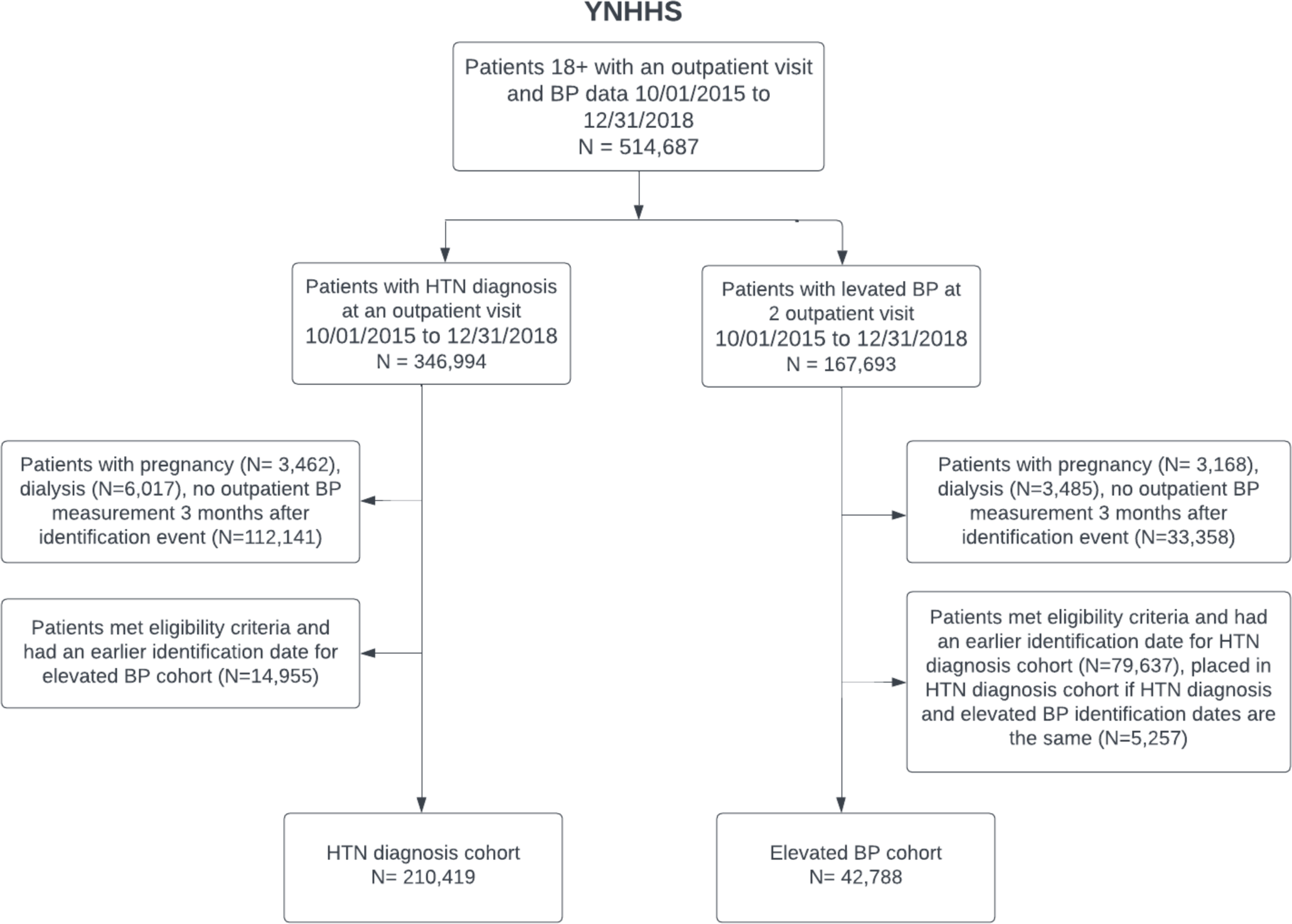
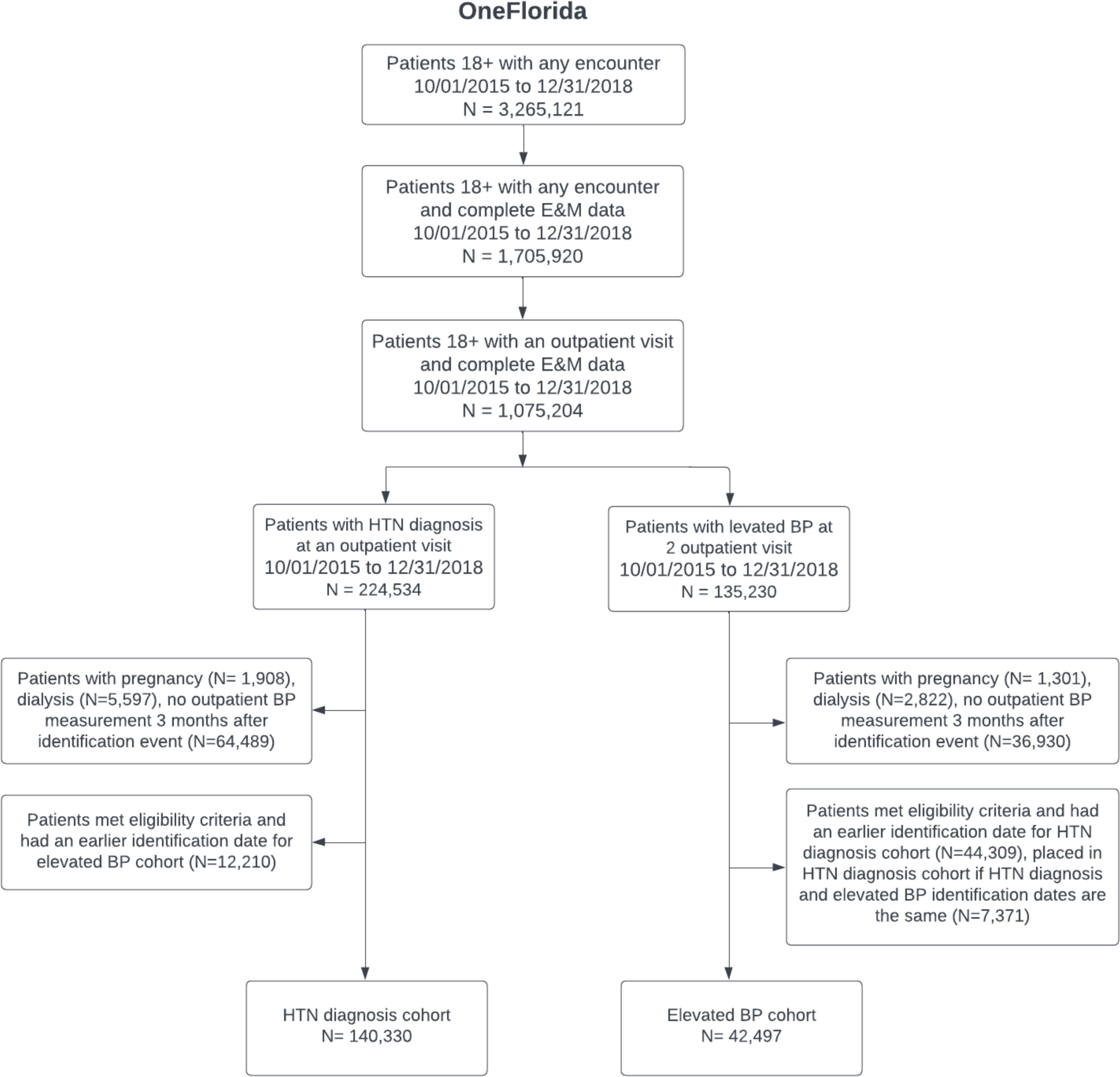
Diagram for study population selection.

### Prevalence and characteristics of uncontrolled hypertension

Using approach 1 as the main analysis, we found uncontrolled hypertension was common and observed among 32.5% of patients at YNHHS and 43.7% of patients at OneFlorida (**Table 1**). The rates of uncontrolled hypertension were similar across age cohorts for OneFlorida (18-44 years: 43.3%; 45-64 years: 43.7%, 65+ years: 43.8%; p=0.15) and trended towards lower rates in older age cohorts for YNHHS (18-44 years: 33.7%, 45-64 years: 33.0%, 65+ years: 31.9%; p<0.001), respectively. Rates of uncontrolled hypertension were similar among women and men at OneFlorida (women: 43.9%, men: 43.6%; p=0.20), but slightly higher for men at YNHHS (women: 31.8%, men: 33.2%; p <0.001). In both YNHHS and OneFlorida, uncontrolled hypertension was disproportionately higher among Black patients when compared with White patients (38.9% versus 31.5%; p<0.01; 49.7% versus 41.2%; p<0.001) and higher among patients with obesity defined as BMI ≥ 30 kg/m^2^ compared to those with BMI < 25 kg/m^2^ (34.1% versus 29.5%; p<0.001; 45.4% versus 41.0%; p<0.001). However, patients with uncontrolled hypertension had fewer comorbidities overall.

**Table 1.**
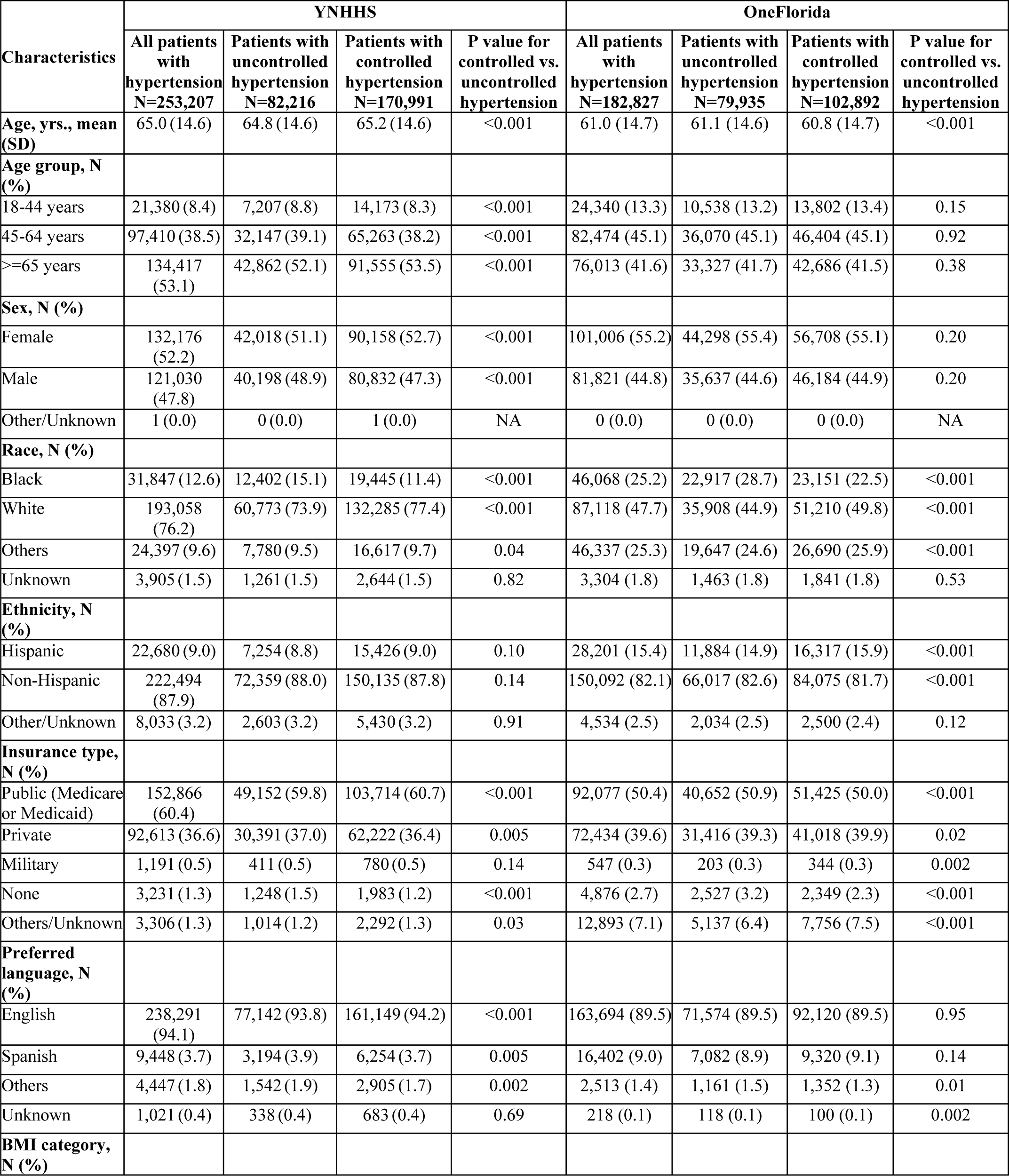
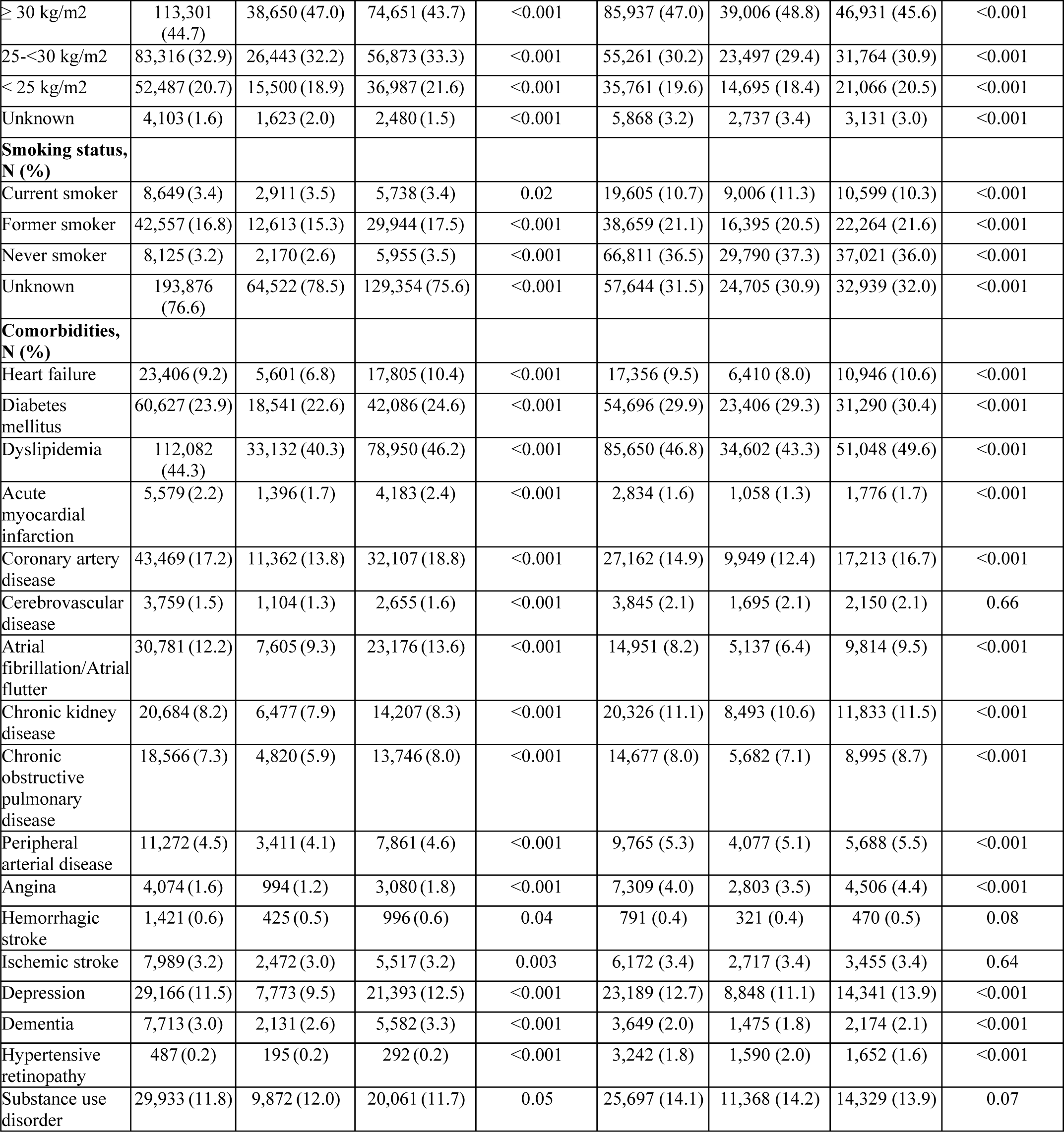
Baseline characteristics of patients with hypertension at the index encounter.

| Characteristics | YNHHS |  |  |  | OneFlorida |  |  |  |
| --- | --- | --- | --- | --- | --- | --- | --- | --- |
|  | All patients with hypertension<br>N=253,207 | Patients with uncontrolled hypertension<br>N=82,216 | Patients with controlled hypertension<br>N=170,991 | P value for controlled vs. uncontrolled hypertension | All patients with hypertension<br>N=182,827 | Patients with uncontrolled hypertension<br>N=79,935 | Patients with controlled hypertension<br>N=102,892 | P value for controlled vs. uncontrolled hypertension |
| **Age, yrs., mean (SD)** | 65.0 (14.6) | 64.8 (14.6) | 65.2 (14.6) | <0.001 | 61.0 (14.7) | 61.1 (14.6) | 60.8 (14.7) | <0.001 |
| **Age group, N (%)** |  |  |  |  |  |  |  |  |
| 18-44 years | 21,380 (8.4) | 7,207 (8.8) | 14,173 (8.3) | <0.001 | 24,340 (13.3) | 10,538 (13.2) | 13,802 (13.4) | 0.15 |
| 45-64 years | 97,410 (38.5) | 32,147 (39.1) | 65,263 (38.2) | <0.001 | 82,474 (45.1) | 36,070 (45.1) | 46,404 (45.1) | 0.92 |
| >=65 years | 134,417 (53.1) | 42,862 (52.1) | 91,555 (53.5) | <0.001 | 76,013 (41.6) | 33,327 (41.7) | 42,686 (41.5) | 0.38 |
| **Sex, N (%)** |  |  |  |  |  |  |  |  |
| Female | 132,176 (52.2) | 42,018 (51.1) | 90,158 (52.7) | <0.001 | 101,006 (55.2) | 44,298 (55.4) | 56,708 (55.1) | 0.20 |
| Male | 121,030 (47.8) | 40,198 (48.9) | 80,832 (47.3) | <0.001 | 81,821 (44.8) | 35,637 (44.6) | 46,184 (44.9) | 0.20 |
| Other/Unknown | 1 (0.0) | 0 (0.0) | 1 (0.0) | NA | 0 (0.0) | 0 (0.0) | 0 (0.0) | NA |
| **Race, N (%)** |  |  |  |  |  |  |  |  |
| Black | 31,847 (12.6) | 12,402 (15.1) | 19,445 (11.4) | <0.001 | 46,068 (25.2) | 22,917 (28.7) | 23,151 (22.5) | <0.001 |
| White | 193,058 (76.2) | 60,773 (73.9) | 132,285 (77.4) | <0.001 | 87,118 (47.7) | 35,908 (44.9) | 51,210 (49.8) | <0.001 |
| Others | 24,397 (9.6) | 7,780 (9.5) | 16,617 (9.7) | 0.04 | 46,337 (25.3) | 19,647 (24.6) | 26,690 (25.9) | <0.001 |
| Unknown | 3,905 (1.5) | 1,261 (1.5) | 2,644 (1.5) | 0.82 | 3,304 (1.8) | 1,463 (1.8) | 1,841 (1.8) | 0.53 |
| **Ethnicity, N (%)** |  |  |  |  |  |  |  |  |
| Hispanic | 22,680 (9.0) | 7,254 (8.8) | 15,426 (9.0) | 0.10 | 28,201 (15.4) | 11,884 (14.9) | 16,317 (15.9) | <0.001 |
| Non-Hispanic | 222,494 (87.9) | 72,359 (88.0) | 150,135 (87.8) | 0.14 | 150,092 (82.1) | 66,017 (82.6) | 84,075 (81.7) | <0.001 |
| Other/Unknown | 8,033 (3.2) | 2,603 (3.2) | 5,430 (3.2) | 0.91 | 4,534 (2.5) | 2,034 (2.5) | 2,500 (2.4) | 0.12 |
| **Insurance type, N (%)** |  |  |  |  |  |  |  |  |
| Public (Medicare or Medicaid) | 152,866 (60.4) | 49,152 (59.8) | 103,714 (60.7) | <0.001 | 92,077 (50.4) | 40,652 (50.9) | 51,425 (50.0) | <0.001 |
| Private | 92,613 (36.6) | 30,391 (37.0) | 62,222 (36.4) | 0.005 | 72,434 (39.6) | 31,416 (39.3) | 41,018 (39.9) | 0.02 |
| Military | 1,191 (0.5) | 411 (0.5) | 780 (0.5) | 0.14 | 547 (0.3) | 203 (0.3) | 344 (0.3) | 0.002 |
| None | 3,231 (1.3) | 1,248 (1.5) | 1,983 (1.2) | <0.001 | 4,876 (2.7) | 2,527 (3.2) | 2,349 (2.3) | <0.001 |
| Others/Unknown | 3,306 (1.3) | 1,014 (1.2) | 2,292 (1.3) | 0.03 | 12,893 (7.1) | 5,137 (6.4) | 7,756 (7.5) | <0.001 |
| **Preferred language, N (%)** |  |  |  |  |  |  |  |  |
| English | 238,291 (94.1) | 77,142 (93.8) | 161,149 (94.2) | <0.001 | 163,694 (89.5) | 71,574 (89.5) | 92,120 (89.5) | 0.95 |
| Spanish | 9,448 (3.7) | 3,194 (3.9) | 6,254 (3.7) | 0.005 | 16,402 (9.0) | 7,082 (8.9) | 9,320 (9.1) | 0.14 |
| Others | 4,447 (1.8) | 1,542 (1.9) | 2,905 (1.7) | 0.002 | 2,513 (1.4) | 1,161 (1.5) | 1,352 (1.3) | 0.01 |
| Unknown | 1,021 (0.4) | 338 (0.4) | 683 (0.4) | 0.69 | 218 (0.1) | 118 (0.1) | 100 (0.1) | 0.002 |
| **BMI category, N (%)** |  |  |  |  |  |  |  |  |
| ≥ 30 kg/m2 | 113,301 (44.7) | 38,650 (47.0) | 74,651 (43.7) | <0.001 | 85,937 (47.0) | 39,006 (48.8) | 46,931 (45.6) | <0.001 |
| 25-<30 kg/m2 | 83,316 (32.9) | 26,443 (32.2) | 56,873 (33.3) | <0.001 | 55,261 (30.2) | 23,497 (29.4) | 31,764 (30.9) | <0.001 |
| < 25 kg/m2 | 52,487 (20.7) | 15,500 (18.9) | 36,987 (21.6) | <0.001 | 35,761 (19.6) | 14,695 (18.4) | 21,066 (20.5) | <0.001 |
| Unknown | 4,103 (1.6) | 1,623 (2.0) | 2,480 (1.5) | <0.001 | 5,868 (3.2) | 2,737 (3.4) | 3,131 (3.0) | <0.001 |
| **Smoking status, N (%)** |  |  |  |  |  |  |  |  |
| Current smoker | 8,649 (3.4) | 2,911 (3.5) | 5,738 (3.4) | 0.02 | 19,605 (10.7) | 9,006 (11.3) | 10,599 (10.3) | <0.001 |
| Former smoker | 42,557 (16.8) | 12,613 (15.3) | 29,944 (17.5) | <0.001 | 38,659 (21.1) | 16,395 (20.5) | 22,264 (21.6) | <0.001 |
| Never smoker | 8,125 (3.2) | 2,170 (2.6) | 5,955 (3.5) | <0.001 | 66,811 (36.5) | 29,790 (37.3) | 37,021 (36.0) | <0.001 |
| Unknown | 193,876 (76.6) | 64,522 (78.5) | 129,354 (75.6) | <0.001 | 57,644 (31.5) | 24,705 (30.9) | 32,939 (32.0) | <0.001 |
| **Comorbidities, N (%)** |  |  |  |  |  |  |  |  |
| Heart failure | 23,406 (9.2) | 5,601 (6.8) | 17,805 (10.4) | <0.001 | 17,356 (9.5) | 6,410 (8.0) | 10,946 (10.6) | <0.001 |
| Diabetes mellitus | 60,627 (23.9) | 18,541 (22.6) | 42,086 (24.6) | <0.001 | 54,696 (29.9) | 23,406 (29.3) | 31,290 (30.4) | <0.001 |
| Dyslipidemia | 112,082 (44.3) | 33,132 (40.3) | 78,950 (46.2) | <0.001 | 85,650 (46.8) | 34,602 (43.3) | 51,048 (49.6) | <0.001 |
| Acute myocardial infarction | 5,579 (2.2) | 1,396 (1.7) | 4,183 (2.4) | <0.001 | 2,834 (1.6) | 1,058 (1.3) | 1,776 (1.7) | <0.001 |
| Coronary artery disease | 43,469 (17.2) | 11,362 (13.8) | 32,107 (18.8) | <0.001 | 27,162 (14.9) | 9,949 (12.4) | 17,213 (16.7) | <0.001 |
| Cerebrovascular disease | 3,759 (1.5) | 1,104 (1.3) | 2,655 (1.6) | <0.001 | 3,845 (2.1) | 1,695 (2.1) | 2,150 (2.1) | 0.66 |
| Atrial fibrillation/Atrial flutter | 30,781 (12.2) | 7,605 (9.3) | 23,176 (13.6) | <0.001 | 14,951 (8.2) | 5,137 (6.4) | 9,814 (9.5) | <0.001 |
| Chronic kidney disease | 20,684 (8.2) | 6,477 (7.9) | 14,207 (8.3) | <0.001 | 20,326 (11.1) | 8,493 (10.6) | 11,833 (11.5) | <0.001 |
| Chronic obstructive pulmonary disease | 18,566 (7.3) | 4,820 (5.9) | 13,746 (8.0) | <0.001 | 14,677 (8.0) | 5,682 (7.1) | 8,995 (8.7) | <0.001 |
| Peripheral arterial disease | 11,272 (4.5) | 3,411 (4.1) | 7,861 (4.6) | <0.001 | 9,765 (5.3) | 4,077 (5.1) | 5,688 (5.5) | <0.001 |
| Angina | 4,074 (1.6) | 994 (1.2) | 3,080 (1.8) | <0.001 | 7,309 (4.0) | 2,803 (3.5) | 4,506 (4.4) | <0.001 |
| Hemorrhagic stroke | 1,421 (0.6) | 425 (0.5) | 996 (0.6) | 0.04 | 791 (0.4) | 321 (0.4) | 470 (0.5) | 0.08 |
| Ischemic stroke | 7,989 (3.2) | 2,472 (3.0) | 5,517 (3.2) | 0.003 | 6,172 (3.4) | 2,717 (3.4) | 3,455 (3.4) | 0.64 |
| Depression | 29,166 (11.5) | 7,773 (9.5) | 21,393 (12.5) | <0.001 | 23,189 (12.7) | 8,848 (11.1) | 14,341 (13.9) | <0.001 |
| Dementia | 7,713 (3.0) | 2,131 (2.6) | 5,582 (3.3) | <0.001 | 3,649 (2.0) | 1,475 (1.8) | 2,174 (2.1) | <0.001 |
| Hypertensive retinopathy | 487 (0.2) | 195 (0.2) | 292 (0.2) | <0.001 | 3,242 (1.8) | 1,590 (2.0) | 1,652 (1.6) | <0.001 |
| Substance use disorder | 29,933 (11.8) | 9,872 (12.0) | 20,061 (11.7) | 0.05 | 25,697 (14.1) | 11,368 (14.2) | 14,329 (13.9) | 0.07 |

### Medication prescription patterns

At YNHHS, 62.1% of patients with hypertension, including 60.7% of those with uncontrolled hypertension and 62.7% of those with controlled hypertension (p=0.56), were not prescribed any antihypertensive drugs in the year prior to the index date (**Table 2**). Among all patients with hypertension, ACEIs or ARBs were prescribed in 19.8% of the patients in the year prior to the index date, followed by beta-blockers (15.3%) and CCBs (11.6%). At OneFlorida, 62.0% of patients with hypertension, including 57.8% of those with uncontrolled hypertension and 65.2% of those with controlled hypertension (p<0.001), were not prescribed any antihypertensive drugs in the year prior to the index date. Among all patients with hypertension, ACEIs or ARBs were prescribed in 22.7% of the patients in the year prior to the index date, followed by CCBs (12.9%) and thiazide or thiazide-like diuretics (12.3%). A total of 5.3% of all patients with hypertension at both YNHHS and OneFlorida sites were prescribed single-pill combination antihypertensive drugs.

**Table 2.** Antihypertensive medication classes prescribed for patients with hypertension in the year prior to the index date.

| Medication class | YNHHS |  |  | OneFlorida |  |  |
| --- | --- | --- | --- | --- | --- | --- |
|  | All patients with hypertension<br>N=253,207 | Patients with uncontrolled hypertension<br>N=82,216 | Patients with controlled hypertension<br>N=170,991 | All patients with hypertension<br>N=182,827 | Patients with uncontrolled hypertension<br>N=79,935 | Patients with controlled hypertension<br>N=102,892 |
| Angiotensin-converting enzyme inhibitor (ACEI) | 27,803 (11.0) | 9,566 (10.7) | 18,237 (10.7) | 26,677 (14.6) | 13,070 (16.4) | 13,607 (13.2) |
| Angiotensin receptor blocker (ARB) | 23,439 (9.3) | 8,610 (10.5) | 14,829 (8.7) | 15,950 (8.7) | 8,324 (10.4) | 7,626 (7.4) |
| ACEI or ARB | 50,246 (19.8) | 17,777 (21.6) | 32,469 (19.0) | 41,528 (22.7) | 20,714 (25.9) | 20,814 (20.2) |
| Calcium channel blocker (CCB) | 29,293 (11.6) | 11,926 (14.5) | 17,367 (10.2) | 23,587 (12.9) | 13,402 (16.8) | 10,185 (9.9) |
| Beta-blocker | 38,757 (15.3) | 12,135 (14.8) | 26,622 (15.6) | 21,901 (12.0) | 10,484 (13.1) | 11,417 (11.1) |
| Thiazide or thiazide-like diuretic | 23,552 (9.3) | 9,059 (11.0) | 14,493 (8.5) | 22,409 (12.3) | 11,749 (14.7) | 10,660 (10.4) |
| Other antihypertensive drug classes | 20,562 (8.1) | 5,922 (7.2) | 14,640 (8.6) | 13,442 (7.4) | 6,430 (8.0) | 7,012 (6.8) |
| Combination antihypertensive drug | 13,530 (5.3) | 4,654 (5.7) | 8,876 (5.2) | 9,722 (5.3) | 4,801 (6.0) | 4,921 (4.8) |
| None | 157,169 (62.1) | 49,942 (60.7) | 107,227 (62.7) | 113,336 (62.0) | 46,232 (57.8) | 67,104 (65.2) |

Similarly, over 50% of patients with hypertension were not prescribed any antihypertensive drugs on the index date. This was consistent across age, sex, and controlled/uncontrolled hypertension subgroups at both YNHHS and OneFlorida sites (**Table 3**). Among patients prescribed at least one antihypertensive drug, 40-50% of patients at YNHHS and 50%-60% of patients at OneFlorida were prescribed one drug class, 20-30% at YNHHS and OneFlorida were prescribed drugs from two drug classes and 10-20% at YNHHS and OneFlorida were prescribed three or more drug classes.

**Table 3.** Number of antihypertensive medication classes prescribed on the index date among patients with hypertension, according to age and sex.

**(A) YNHHS**
| Number of medication classes | Patients with uncontrolled hypertension |  |  |  |  |  | Patients with controlled hypertension |  |  |  |  |  |
| --- | --- | --- | --- | --- | --- | --- | --- | --- | --- | --- | --- | --- |
|  | Men |  |  | Women |  |  | Men |  |  | Women |  |  |
|  | 18-44 years | 45-64 years | 65+ years | 18-44 years | 45-64 years | 65+ years | 18-44 years | 45-64 years | 65+ years | 18-44 years | 45-64 years | 65+ years |
| 0 | 2,556 (60.9) | 9,665 (55.3) | 11,117 (60.0) | 1,939 (64.5) | 8,829 (60.2) | 14,392 (59.1) | 5,037 (73.1) | 19,538 (62.0) | 25,857 (61.0) | 5,527 (75.9) | 22,765 (67.5) | 30,870 (62.8) |
| 1 | 938 (22.3) | 3,948 (22.6) | 3,869 (20.9) | 643 (21.4) | 3,107 (21.2) | 5,088 (20.9) | 1,146 (16.6) | 6,851 (21.7) | 9,462 (22.3) | 1,212 (16.7) | 6,467 (19.2) | 10,365 (21.1) |
| 2 | 492 (11.7) | 2,461 (14.1) | 2,202 (11.9) | 287 (9.5) | 1,777 (12.1) | 3,113 (12.8) | 505 (7.3) | 3,630 (11.5) | 4,723 (11.1) | 408 (5.6) | 3,308 (9.8) | 5,372 (10.9) |
| ≥3 | 214 (5.1) | 1,401 (8.0) | 1,335 (7.2) | 138 (4.6) | 959 (6.5) | 1,746 (7.2) | 207 (3.0) | 1,512 (4.8) | 2,364 (5.6) | 131 (1.8) | 1,191 (3.5) | 2,542 (5.2) |

**(B) OneFlorida**
| Number of medication classes | Patients with uncontrolled hypertension |  |  |  |  |  | Patients with controlled hypertension |  |  |  |  |  |
| --- | --- | --- | --- | --- | --- | --- | --- | --- | --- | --- | --- | --- |
|  | Men |  |  | Women |  |  | Men |  |  | Women |  |  |
|  | 18-44 years | 45-64 years | 65+ years | 18-44 years | 45-64 years | 65+ years | 18-44 years | 45-64 years | 65+ years | 18-44 years | 45-64 years | 65+ years |
| 0 | 3,370 (67.1) | 10,647 (64.2) | 10,453 (74.5) | 3,646 (66.1) | 12,052 (61.8) | 13,462 (69.8) | 4,667 (78.9) | 14,573 (72.7) | 15,998 (79.1) | 6,094 (77.3) | 18,850 (71.5) | 17,218 (76.7) |
| 1 | 1,018 (20.3) | 3,049 (18.4) | 1,979 (14.1) | 1,058 (19.2) | 3,703 (19.0) | 3,010 (15.6) | 834 (14.1) | 3,164 (15.8) | 2,507 (12.4) | 1,218 (15.4) | 4,229 (16) | 2,981 (13.3) |
| 2 | 420 (8.4) | 1,784 (10.8) | 1,016 (7.2) | 544 (9.9) | 2,302 (11.8) | 1,710 (8.9) | 308 (5.2) | 1,640 (8.2) | 1,189 (5.9) | 430 (5.5) | 2,353 (8.9) | 1,563 (7.0) |
| ≥3 | 212 (4.2) | 1,097 (6.6) | 592 (4.2) | 270 (4.9) | 1,436 (7.4) | 1,105 (5.7) | 105 (1.8) | 665 (3.3) | 534 (2.6) | 146 (1.9) | 930 (3.5) | 696 (3.1) |

Among adults prescribed one antihypertensive medication class on the index date, ACEI or ARBs was the most prescribed class at both YNHHS and OneFlorida (34.3% at YNHHS and 40.5% at OneFlorida; **Table 4**). For YNHHS, the second most prescribed medication class was beta-blockers (28.4%) followed by CCBs (18.9%). For OneFlorida, the second most prescribed medication class was CCBs (19.8%) followed by beta blockers (18.8%). Among adults prescribed two antihypertensive drug classes, ACEI or ARB and thiazide diuretic were most common (25.8% at YNHHS and 33.1% at OneFlorida). Among patients using three or more antihypertensive drug classes, ACEI or ARB, CCB and thiazide diuretic were most common (16.9% at YNHHS and 20.9% at OneFlorida).

**Table 4.** Top three commonly prescribed antihypertensive medication classes on the index date among treated patients with hypertension.

**(A) YNHHS**
|  | **All patients with hypertension**<br>**N=253,207** | **Patients with uncontrolled hypertension**<br>**N=82,216** | **Patients with controlled hypertension**<br>**N=170,991** |
| --- | --- | --- | --- |
| **Among adults using one medication class** |  |  |  |
| ACEI or ARB* | 18,216 (34.3) | 6,353 (36.1) | 11,863 (33.4) |
| Beta blocker | 15,086 (28.4) | 4,191 (23.8) | 10,895 (30.7) |
| CCB | 10,037 (18.9) | 4,077 (23.2) | 5,960 (16.8) |
| Total | 53,097 | 17,593 | 35,504 |
| **Among adults using two medication classes** |  |  |  |
| ACEI or ARB and Thiazide diuretic | 7,307 (25.8) | 2,729 (26.4) | 4,578 (25.5) |
| ACEI or ARB and Beta blocker | 3,421 (18.5) | 1,698 (16.4) | 3,541 (19.7) |
| ACEI or ARB and CCB | 3,142 (16.2) | 2,016 (19.5) | 2,557 (14.2) |
| Total | 28,278 | 10,332 | 17,946 |
| **Among adults using three or more medication classes** |  |  |  |
| ACEI or ARB and CCB and Thiazide diuretic | 2,321 (16.9) | 1,140 (19.7) | 1,181 (14.9) |
| ACEI or ARB and Beta Blocker and Thiazide diuretic | 1,913 (13.9) | 1,137 (14.3) | 776 (13.4) |
| ACEI or ARB and Beta Blocker and CCB | 1,722 (12.5) | 786 (13.6) | 936 (11.8) |
| Total | 13,740 | 5,793 | 7,947 |

**(B) OneFlorida**
|  | **All patients with hypertension**<br>**N=182,827** | **Patients with uncontrolled hypertension**<br>**N=79,935** | **Patients with controlled hypertension**<br>**N=102,892** |
| --- | --- | --- | --- |
| **Among adults using one medication class** |  |  |  |
| ACEI or ARB | 11,638 (40.5) | 5,597 (40.5) | 6,041 (40.5) |
| CCB | 5,703 (19.8) | 3,140 (22.7) | 2,563 (17.2) |
| Beta Blocker | 5,406 (18.8) | 2,292 (16.6) | 3,114 (20.9) |
| Total | 28,750 (100) | 13,817 (100) | 14,933 (100) |
| **Among adults using two medication classes** |  |  |  |
| ACEI or ARB and Thiazide diuretic | 5,046 (33.1) | 2,443 (31.4) | 2,603 (34.8) |
| ACEI or ARB and CCB | 2,753 (18) | 1,600 (20.6) | 1,153 (15.4) |
| ACEI or ARB and Beta Blocker | 2,182 (14.3) | 1,041 (13.4) | 1,141 (15.2) |
| Total | 15,259 (100) | 7,776 (100) | 7,483 (100) |
| **Among adults using three or more medication classes** |  |  |  |
| ACEI or ARB and CCB and Thiazide diuretic | 1,629 (20.9) | 1,050 (22.3) | 579 (18.8) |
| ACEI or ARB and Beta Blocker and Thiazide diuretic | 985 (12.6) | 530 (11.2) | 455 (14.8) |
| ACEI or ARB and Beta Blocker and CCB | 763 (9.8) | 460 (9.8) | 303 (9.9) |
| Total | 7,788 (100) | 4,712 (100) | 3,076 (100) |

\*ACEI: Angiotensin-converting enzyme inhibitor; ARB: Angiotensin receptor blocker; CCB: Calcium channel blocker.

### Short-term and long-term outcomes

Overall, the composite of death and CVD event rates among patients with hypertension at 3, 6, 12 and 24 months were 3.3%, 5.4%, 5.4% and 8.6% at YNHHS; the rates were 1.9%, 2.9%, 4.3% and 6.0% at OneFlorida (**Table 5**). The proportion of patients who had ED or inpatient visits for any cause at 3, 6, 12 and 24 months were 12.8%, 19.8%, 19.9% and 29.7% at YNHHS; the proportions were 10.7%, 15.8%, 22.7% and 29.6% at OneFlorida. The proportion of patients who had ambulatory visits for any cause at 3, 6, 12 and 24 months were 75.4%, 87.0%, 88.7% and 95.2% at YNHHS; the proportions were 50.8%, 68.1%, 79.6% and 84.6% at OneFlorida. Patients with controlled and uncontrolled hypertension had similar rates of short-term (at 3 and 6 months) and long-term (at 12 and 24 months) clinical outcomes and healthcare utilizations.

**Table 5.** Rates of death, non-fatal CVD events, and healthcare utilization, among patients with uncontrolled and controlled hypertension at two health systems at 3, 6, 12, 24 months after the index date.

|  | YNHHS |  |  |  | OneFlorida |  |  |  |
| --- | --- | --- | --- | --- | --- | --- | --- | --- |
|  | Event rate at 3 months (%) | Event rate at 6 months (%) | Event rate at 12 months (%) | Event rate at 24 months (%) | Event rate at 3 months (%) | Event rate at 6 months (%) | Event rate at 12 months (%) | Event rate at 24 months (%) |
| **All clinical outcomes\*** |  |  |  |  |  |  |  |  |
| All patients | 3.3 | 5.4 | 5.4 | 8.6 | 1.9 | 2.9 | 4.3 | 6.0 |
| Uncontrolled | 2.7 | 4.5 | 4.5 | 7.6 | 1.6 | 2.4 | 3.7 | 5.4 |
| Controlled | 3.6 | 5.8 | 5.8 | 9.1 | 2.1 | 3.2 | 4.7 | 6.5 |
| **Death** |  |  |  |  |  |  |  |  |
| All patients | 1.2 | 2.2 | 2.2 | 3.7 | 1.4 | 2.3 | 3.7 | 5.7 |
| Uncontrolled | 0.8 | 1.5 | 1.6 | 2.7 | 1.2 | 2.0 | 3.2 | 5.0 |
| Controlled | 1.4 | 2.4 | 2.5 | 4.1 | 1.6 | 2.6 | 4.1 | 6.1 |
| **CVD events** |  |  |  |  |  |  |  |  |
| All patients | 2.6 | 4.1 | 4.1 | 6.5 | 2.1 | 3.3 | 5.4 | 7.8 |
| Uncontrolled | 2.2 | 3.7 | 3.7 | 6.1 | 1.9 | 3.2 | 5.3 | 7.8 |
| Controlled | 2.8 | 4.3 | 4.4 | 6.7 | 2.2 | 3.5 | 5.5 | 7.8 |
| **ED and/or inpatient visit for any cause** |  |  |  |  |  |  |  |  |
| All patients | 12.8 | 19.8 | 19.9 | 29.7 | 10.7 | 15.8 | 22.7 | 29.6 |
| Uncontrolled | 11.8 | 18.4 | 18.6 | 28.2 | 10.0 | 15.0 | 21.8 | 28.6 |
| Controlled | 13.3 | 20.4 | 20.5 | 30.4 | 11.2 | 16.5 | 23.5 | 30.3 |
| **Ambulatory visit for any cause** |  |  |  |  |  |  |  |  |
| All patients | 75.4 | 87.0 | 88.7 | 95.2 | 50.8 | 68.1 | 79.6 | 84.6 |
| Uncontrolled | 74.3 | 86.5 | 88.2 | 95.0 | 49.6 | 66.9 | 78.6 | 84.0 |
| Controlled | 75.9 | 87.3 | 89.0 | 95.3 | 51.8 | 69.1 | 80.4 | 85.1 |

\*All clinical outcomes include the composite of death and non-fatal CVD events.

The results of sensitivity analysis using approach 2 where we defined controlled and uncontrolled hypertension based on a single BP measurement at the index visit were reported in **Supplemental Tables S6-S10**. The sensitivity analysis showed results consistent with the main analysis.

## DISCUSSION

Our study applied two computable definitions to EHR data from two large clinical research networks, YNHHS and OneFlorida, to identify and characterize patient populations with controlled and uncontrolled hypertension. The two computable definitions generated consistent results. Approximately 30-40% of hypertensive patients receiving ambulatory care within both health system networks have uncontrolled hypertension, of whom 60% were untreated. We were also able to characterize short-term and long-term outcomes among patients with both controlled and uncontrolled hypertension. These findings lay a foundation for more sophisticated analyses to assess the quality of care and outcomes for patients with hypertension in future studies.

A strength of this study was the successful use of a decentralized model for clinical research. Both YNHHS and OneFlorida retained their data behind their individual firewalls, but data were managed using common definitions and data models that enabled harmonized research using federated analytics. Conducting clinical research using federated models enables aggregation of observations across multiple health systems, thereby examining a much larger and diverse population size of patients than when using data from a single health system. The consistent overall results that we found across both YNHHS and OneFlorida suggest that a reusable infrastructure can be created for digital population health surveillance and identification of people with hypertension who would benefit from more aggressive management.

Several challenges were encountered during the study, as well as insights that have led us to conclude that they are all addressable. An overall challenge was accurately defining and identifying a condition-specific population, in this case patients with uncontrolled hypertension. To use EHR data to perform high-quality clinical research, construction of accurate patient cohorts is vital. This is particularly important for uncontrolled hypertension, for which there is no specific diagnostic code and identification usually requires many observations over time. Clinical guidelines have established a fundamental definition of uncontrolled hypertension based on BP thresholds.^3, 9^ For instance, the Joint National Committee on Prevention, Detection, Evaluation, and Treatment of High Blood Pressure (JNC-8) defined uncontrolled hypertension as a BP level greater than or equal to 140/90 mmHg. In contrast, the 2017 hypertension guideline recommended a lower BP threshold for defining uncontrolled hypertension, specifically, a BP level greater than or equal to 130/80 mmHg. This study represents additional work to develop computable phenotypes for uncontrolled hypertension based on ICD10-CM codes, BP measurements, and using common data models (in this case the PCORnet common data model) for use in clinical research and practice.

Importantly, previous studies have shown that diagnosis codes used in isolation generally do not have sufficient accuracy for cohort identification. Even for a straightforward diagnosis such as hypertension, approximately 30% of the people identified with hypertension by BP measurements recorded in the EHR were missing the associated diagnostic code.^20, 21^ We found a similar proportion of hypertensive patients did not have associated diagnostic code. One solution to improve the robustness of results, as we showed in this study, is to develop different operational definitions of uncontrolled hypertension and evaluate how these definitions may influence cohort identification. With the increasing emphasis on ambulatory and home BP monitoring,^22, 23^ additional data sources may be available to better understand the management of hypertension when these data are integrated with the EHR.

Second, using health system data to classify antihypertensive medications and examine patterns of medication prescription has challenges. This is because many medications have multiple indications and dosage forms, and the existing therapeutic classification systems generally group medications in ways that may only partially correlate with intended use. For example, timolol is a beta-blocker that has both oral and ophthalmic dosage forms. The oral form is used to treat hypertension, whereas the ophthalmic form is used to treat glaucoma.^24, 25^ Therefore, just the presence of a drug entity in the prescription records may not be sufficient to accurately classify medications being used for hypertension treatment. A solution is to use a set of standardized drug codes and names for use in querying EHR data for antihypertensive medication prescriptions.^17^ This approach allowed us to properly identify antihypertensive medications, assign each medication to a medication class, and apply consistent definitions across multiple health systems. Of note, we found over 50% of patients with controlled hypertension were not on antihypertensive medications. Likely, these individuals were able to achieve their BP goals through non-pharmacologic means. Lifestyle modifications, such as adopting a healthy diet, engaging in regular physical activity, and reducing stress, have been shown to have a positive impact on BP management. It is also possible that these patients were effectively treating and managing underlying medical conditions that contribute to elevated BP, such as obstructive sleep apnea, chronic kidney disease, or hormonal disorders. In addition, the distribution of prescribed antihypertensive medications varied across health systems, as the specific selection of medication depends on multiple factors. For instance, diuretics may be favored for hypertensive patients experiencing fluid retention, while beta-blockers might be more suitable for those with a history of heart disease or arrhythmia. Similarly, hypertensive patients with diabetes or chronic kidney disease may prefer ACE inhibitors or ARBs due to their additional renal protective effects. Moreover, the choice of antihypertensive medication can be influenced by the preferences and familiarity of the prescribing physician with different medication classes. Some physicians may possess greater expertise in certain medications or prefer those with fewer side effects and better tolerability profiles.

Third, there were pros and cons of using the primary discharge diagnosis codes versus secondary diagnosis codes to identify the outcomes of interest across health systems. Using primary discharge diagnosis codes for hospitalizations for CVD events like stroke may be less likely to have misclassification than codes from ambulatory visits. However, some events may be missed by reliance solely on primary diagnosis codes, particularly when there are concurrent diagnoses. On the other hand, including secondary diagnoses may lead to greater capture of events, but it may lead to too much noise resulting from the inability to distinguish patients with acute strokes from those with history of prior stroke. The approach we used in this study was to include only acute event codes – whether or not they were in the primary diagnosis position – for outcome ascertainment. Another common solution for improving accuracy of outcome ascertainment is to validate the diagnosis codes against manual chart review, as showed in prior EHR studies.^26^ While our study did not perform chart review due to the limited scope of work, comparing the diagnostic codes or algorithms with clinician review of EHRs to determine extent of concordance between codes and clinical judgement may be necessary to evaluate and improve the validity of codes or algorithms. There is also a critical need to ensure that these methods are consistent across different sites within the distributed research model. Of note, it is crucial to recognize that the present study adopts a descriptive design and does not aim to evaluate the association between hypertension control and clinical outcomes. As a result, the controlled and uncontrolled hypertension groups may exhibit different demographic or clinical characteristics that were not accounted for in the outcome analysis. The controlled hypertension group might have been composed of individuals who were more proactive in managing their condition and adhering to treatment regimens. This self-selection bias could indicate that these patients were generally more engaged in their health, leading to higher healthcare utilization and subsequent identification of clinical events. Another plausible explanation is that patients with more severe or complicated health conditions were prioritized for intensive treatment and achieved controlled hypertension. Therefore, the higher clinical outcomes observed in this group could be attributed to their underlying medical complexity rather than the effect of blood pressure control itself. Finally, it is possible that unmeasured or unknown confounders influenced both the choice of treatment strategy and the clinical outcomes.

### Limitations

There are several limitations in this study. First, there may be variations in methods and devices used to measure BP across and within the two health systems. Measurement of BP in a clinical practice setting may not mirror that of a trial or be performed per best practices. Second, we only used prescribing data to evaluate antihypertensive medications and do not have information on whether the prescriptions were filled or taken by the patients. Third, we used ED or inpatient encounters in the EHR to define clinical outcomes, which presumes that patients were hospitalized at the given health system of interest. For acute events such as myocardial infraction and stroke, patients are often taken by ambulance to the nearest hospital, which may not always be within the YNHHS or OneFlorida network. Thus, there may be incomplete ascertainment of acute events in EHRs. There is also a possibility of misclassification of events, as we employed diagnosis codes in any position and encompassed a wide range of outcomes in our analysis. Fourth, we performed only simple descriptive analyses to evaluate clinical outcomes in this study and did not apply risk adjustment. Fifth, we defined patients’ comorbid conditions by utilizing ICD-10 codes that were recorded within the past year. The purpose of this approach was to capture the patient’s current clinical status. However, we acknowledge that this method may overlook comorbidities that have not been actively managed or diagnosed within the past year, yet still hold the potential to pose future cardiovascular risk. Finally, our findings may not be generalizable to other health systems. This may be due to data limitations (e.g., lack of a common data model) or differences in population and practice patterns. This has potential implications for the scalability of a real-world hypertension surveillance program.

### Conclusions

Real-world data collected during routine care hold great promise for use in clinical research. The current study demonstrates the potential for leveraging EHR data and using computable definitions to conduct digital population surveillance for hypertension management and identify target patients with uncontrolled hypertension who may benefit from additional treatment. This study also describes challenges inherent in performing studies using health system data and strategies to overcome these challenges. These findings provide insights into using real-world data to generate high-quality real-world evidence that can be used to support decisions by regulators, clinicians, and patients.

## Data Availability

The summary data for this manuscript is available upon request to corresponding authors.

## Access to Data Statement

Drs. Lu, Keeley, and Ross had full access to all the data in the study and take responsibility for the integrity of the data and the accuracy of the data analysis.

## Non-standard Abbreviations and Acronyms

ACEI: Angiotensin-converting enzyme inhibitor
AMI: Acute myocardial infarction
ARB: Angiotensin receptor blocker
BP: Blood pressure
CCB: Calcium channel blocker
CVD: Cardiovascular disease
DBP: Diastolic blood pressure
ED: Emergency department
EHR: Electronic health record
ICD-10-CM: International Classification of Diseases-10th Edition-Clinical Modification
NESTcc: National Evaluation System for health Technology Coordinating Center
PCORnet: National Patient-Centered Clinical Research Network
SBP: Systolic blood pressure
SD: Standard deviation
STROBE: Strengthening the Reporting of Observational Studies in Epidemiology
YNHHS: Yale-New Haven Health System

## References

1. Adams JM and Wright JS. A National Commitment to Improve the Care of Patients With Hypertension in the US. JAMA. 2020.

2. Muntner P, Hardy ST, Fine LJ, Jaeger BC, Wozniak G, Levitan EB and Colantonio LD. Trends in Blood Pressure Control Among US Adults With Hypertension, 1999-2000 to 2017-2018. JAMA. 2020;324:1190–1200.

3. Whelton PK, Carey RM, Aronow WS, Casey DE, Jr., Collins KJ, Dennison Himmelfarb C, DePalma SM, Gidding S, Jamerson KA, Jones DW, MacLaughlin EJ, Muntner P, Ovbiagele B, Smith SC, Jr., Spencer CC, Stafford RS, Taler SJ, Thomas RJ, Williams KA, Sr., Williamson JD and Wright JT, Jr. 2017 ACC/AHA/AAPA/ABC/ACPM/AGS/APhA/ASH/ASPC/NMA/PCNA Guideline for the Prevention, Detection, Evaluation, and Management of High Blood Pressure in Adults: Executive Summary: A Report of the American College of Cardiology/American Heart Association Task Force on Clinical Practice Guidelines. Circulation. 2018;138:e426–e483.

4. Jollis JG, Ancukiewicz M, DeLong ER, Pryor DB, Muhlbaier LH and Mark DB. Discordance of databases designed for claims payment versus clinical information systems. Implications for outcomes research. Ann Intern Med. 1993;119:844–50.

5. Hartzema AG, Racoosin JA, MaCurdy TE, Gibbs JM and Kelman JA. Utilizing Medicare claims data for real-time drug safety evaluations:is it feasible? Pharmacoepidemiol Drug Saf. 2011;20:684–8.

6. Kohane IS. HEALTH CARE POLICY. Ten things we have to do to achieve precision medicine. Science. 2015;349:37–8.

7. Weber GM, Mandl KD and Kohane IS. Finding the missing link for big biomedical data. JAMA. 2014;311:2479–80.

8. Wei WQ and Denny JC. Extracting research-quality phenotypes from electronic health records to support precision medicine. Genome Med. 2015;7:41.

9. James PA, Oparil S, Carter BL, Cushman WC, Dennison-Himmelfarb C, Handler J, Lackland DT, LeFevre ML, MacKenzie TD, Ogedegbe O, Smith SC, Jr., Svetkey LP, Taler SJ, Townsend RR, Wright JT, Jr., Narva AS and Ortiz E. 2014 evidence-based guideline for the management of high blood pressure in adults: report from the panel members appointed to the Eighth Joint National Committee (JNC 8). JAMA. 2014;311:507–20.

10. Shuren J and Califf RM. Need for a national evaluation system for health technology. JAMA. 2016;316:1153–1154.

11. Kandzari DE, Bohm M, Mahfoud F, Townsend RR, Weber MA, Pocock S, Tsioufis K, Tousoulis D, Choi JW, East C, Brar S, Cohen SA, Fahy M, Pilcher G, Kario K and Investigators SH-OMT. Effect of renal denervation on blood pressure in the presence of antihypertensive drugs: 6-month efficacy and safety results from the SPYRAL HTN-ON MED proof-of-concept randomised trial. Lancet. 2018;391:2346–2355.

12. Bohm M, Kario K, Kandzari DE, Mahfoud F, Weber MA, Schmieder RE, Tsioufis K, Pocock S, Konstantinidis D, Choi JW, East C, Lee DP, Ma A, Ewen S, Cohen DL, Wilensky R, Devireddy CM, Lea J, Schmid A, Weil J, Agdirlioglu T, Reedus D, Jefferson BK, Reyes D, D’Souza R, Sharp ASP, Sharif F, Fahy M, DeBruin V, Cohen SA, Brar S, Townsend RR and Investigators SH-OMP. Efficacy of catheter-based renal denervation in the absence of antihypertensive medications (SPYRAL HTN-OFF MED Pivotal): a multicentre, randomised, sham-controlled trial. Lancet. 2020;395:1444–1451.

13. Schulz WL, Durant TJ, Torre Jr CJ, Hsiao AL and Krumholz HM. Agile health care analytics: enabling real-time disease surveillance with a computational health platform. J Med Internet Res. 2020;22:e18707.

14. McPadden J, Durant TJ, Bunch DR, Coppi A, Price N, Rodgerson K, Torre Jr CJ, Byron W, Hsiao AL and Krumholz HM. Health care and precision medicine research: analysis of a scalable data science platform. J Med Internet Res. 2019;21:e13043.

15. Teixeira PL, Wei WQ, Cronin RM, Mo H, VanHouten JP, Carroll RJ, LaRose E, Bastarache LA, Rosenbloom ST, Edwards TL, Roden DM, Lasko TA, Dart RA, Nikolai AM, Peissig PL and Denny JC. Evaluating electronic health record data sources and algorithmic approaches to identify hypertensive individuals. J Am Med Inform Assoc. 2017;24:162–171.

16. Peng M, Chen G, Kaplan GG, Lix LM, Drummond N, Lucyk K, Garies S, Lowerison M, Weibe S and Quan H. Methods of defining hypertension in electronic medical records: validation against national survey data. J Public Health (Oxf). 2016;38:e392–e399.

17. McDonough CW, Smith SM, Cooper-DeHoff RM and Hogan WR. Optimizing Antihypertensive Medication Classification in Electronic Health Record-Based Data: Classification System Development and Methodological Comparison. JMIR Med Inform. 2020;8:e14777.

18. Suchard MA, Schuemie MJ, Krumholz HM, You SC, Chen R, Pratt N, Reich CG, Duke J, Madigan D, Hripcsak G and Ryan PB. Comprehensive comparative effectiveness and safety of first-line antihypertensive drug classes: a systematic, multinational, large-scale analysis. Lancet. 2019;394:1816–1826.

19. Fleurence RL, Blake K and Shuren J. The future of registries in the era of real-world evidence for medical devices. JAMA cardiology. 2019;4:197–198.

20. Smith SM, McAuliffe K, Hall JM, McDonough CW, Gurka MJ, Robinson TO, Sacco RL, Pepine C, Shenkman E and Cooper-DeHoff RM. Hypertension in Florida: Data From the OneFlorida Clinical Data Research Network. Prev Chronic Dis. 2018;15:E27.

21. Banerjee D, Chung S, Wong EC, Wang EJ, Stafford RS and Palaniappan LP. Underdiagnosis of hypertension using electronic health records. Am J Hypertens. 2012;25:97–102.

22. Krist AH, Davidson KW, Mangione CM, Cabana M, Caughey AB, Davis EM, Donahue KE, Doubeni CA, Kubik M and Li L. Screening for hypertension in adults: US Preventive Services Task Force reaffirmation recommendation statement. JAMA. 2021;325:1650–1656.

23. Muntner P, Shimbo D, Carey RM, Charleston JB, Gaillard T, Misra S, Myers MG, Ogedegbe G, Schwartz JE and Townsend RR. Measurement of blood pressure in humans: a scientific statement from the American Heart Association. Hypertension. 2019;73:e35–e66.

24. Frampton JE and Perry CM. Topical dorzolamide 2%/timolol 0.5% ophthalmic solution: a review of its use in the treatment of glaucoma and ocular hypertension. Drugs Aging. 2006;23:977–95.

25. Harris FJ, Tonkin M, Pratt C, DeMaria AN, Amsterdam EA and Mason DT. Short- and long-term therapy of mild essential hypertension with timolol. Clin Pharmacol Ther. 1981;30:765–72.

26. Saczynski JS, Andrade SE, Harrold LR, Tjia J, Cutrona SL, Dodd KS, Goldberg RJ and Gurwitz JH. A systematic review of validated methods for identifying heart failure using administrative data. Pharmacoepidemiol Drug Saf. 2012;21 Suppl 1:129–40.

